# ModInterv COVID-19: An online platform to monitor the evolution of epidemic curves

**DOI:** 10.1101/2022.01.31.22270192

**Authors:** Arthur A. Brum, Giovani L. Vasconcelos, Gerson C. Duarte-Filho, Raydonal Ospina, Francisco A. G. Almeida, Antônio M. S. Macêdo

## Abstract

We present the software `ModInterv` as an informatics tool to monitor, in an automated and user-friendly manner, the evolution and trend of COVID-19 epidemic curves, both for cases and deaths. The `ModInterv` software uses parametric generalized growth models, together with LOWESS regression analysis, to fit epidemic curves with multiple waves of infections for countries around the world as well as for states and cities in Brazil and the USA. The software automatically accesses publicly available COVID-19 databases maintained by the Johns Hopkins University (for countries as well as states and cities in the USA) and the Federal University of Viçosa (for states and cities in Brazil). The richness of the implemented models lies in the possibility of quantitatively and reliably detecting the distinct acceleration regimes of the disease. We describe the backend structure of software as well as its practical use. The software helps the user not only to understand the current stage of the epidemic in a chosen location but also to make short term predictions as to how the curves may evolve. The app is freely available on the internet (http://fisica.ufpr.br/modinterv), thus making a sophisticated mathematical analysis of epidemic data readily accessible to any interested user.

## 1. Introduction

The complex evolution pattern of an epidemic outbreak, such as the ongoing COVID-19 pandemic, requires a dynamic response of public health policies based on reliable factual evidence and accurate information analysis. Generally speaking, one can identify different stages of response to an outbreak, ranging from the detection of the first cases, involving surveillance systems and especially qualitative measures of risk assessment, followed by the assessment of the dynamics of transmission in the intervention phases, where more complex analyses are required to inform and guide the authorities as to the adoption of appropriate non-pharmacological interventions (e.g., intermittent lockdowns) and, if available, pharmacological measures as well (e.g., vaccination strategies), up to the registration of the last cases that recover or dies (assuming the disease does not become endemic). Finally, a post-intervention stage emerges, where lessons learned can help and improve protocols and preparation for a possible next epidemic.

An important feature of the modern response to epidemics is the increasing focus on exploiting all available data, including geo-referenced information of specific population groups, such as countries, cities, and even smaller administrative units (e.g., neighborhoods or census sectors). Taking into account this wealth of information can help in a macro view of the situation, enabling rapid responses and evidence-based decision making [4, 7]. The use of data and modeling techniques, together with information tools that range from data collection at service points to the generation of informative situational reports (apps, dashboards, tweets, etc.) [28, 3], allows for a better dissemination of reliable information and contributes to an accurate public perception of the epidemic situation.

From this perspective, we can obtain important insights about the evolution of an outbreak in a given population group by analyzing the corresponding epidemic curves, as represented by either the cumulative or the daily number of cases or deaths as a function of time [14, 13]. Epidemic curves are useful in many aspects because they provide a simple visual outline of demographic dynamics, which can be used to assess the growth or decline of an outbreak [19] as well as to assess the effect of intervention measures. In addition, epidemic curves form the raw material used by a wide range of modeling techniques for monitoring and forecasting epidemics [26, 25, 18, 27]. Having good mathematical models to describe the empirical data is a necessary condition for this endeavor. In this context, phenomenological growth models are an important tool in mathematical epidemiology [5, 26, 25], because contrary to other epidemiological models, such as compartment models [15, 10] and agent-based models [16, 21], growth models are mathematically more tractable, numerically easier to implement, and often admit an analytic solution.

In this paper, we present an automated software application, called `ModInterv`, that enables the user to monitor the COVID-19 epidemic curves of cases and deaths for different countries around the world as well as for states and cities in Brazil and the USA. The app implements a general class of generalized logistic growth models with time-dependent parameters to fit the epidemic curves for the selected location. As most places (countries, states, and cities) around the world have exhibited multiple waves of COVID-19 infections, the app `ModInterv` was developed with the specific aim of automatically analyzing epidemic curves with an arbitrary, *a priori* unknown number *N ≥*1 of waves.

Once the user chooses the type of data (i.e., cases or deaths) and the desired location, the app first decides, based on a LOWESS regression algorithm, the number *N* of waves present in the chosen data. After *N* has been determined, the software fits the raw empirical data with a *N* -wave growth model (to be discussed below) and from the bestfit model provides the user with relevant information about the epidemic evolution in the chosen location. The app displays output plots with the theoretical cumulative and daily curves superimposed with the empirical data, where both the starting and peak dates for each wave are given. Also, special marks (colored vertical lines) are drawn in the output plot to indicate the different acceleration regimes during each epidemic wave. Furthermore, by comparing the location of the last data point in relation to these special points, the app informs the user the current stage of the epidemic, according to a refined classification scheme that considers not only the curve acceleration but also its third and fourth derivatives, known respectively as the jerk (or jolt) and the jounce (or snap). The app also allows the user to perform additional analyses of the fitted results by clicking on extra check boxes and moving certain sliding bars.

The mathematical analysis provided by our application can be useful to public health authorities, not only because it indicates the current dynamical stage of the epidemic in a given place and identifies its dynamical regimes up to present time but also because it can help to predict its likely evolution in the near future. This information, combined with other analyses, can in turn help the authorities in their decision making process regarding, say, the adoption or relaxation of non-pharmacological interventions [24]. The type of analysis provided by the `ModInterv` may also be of value to other mathematical epidemiology research groups, as it allows for an easy and reliable determination of important points (e.g., peaks and troughs) of a multiwave epidemic curve—an information that is relevant when studying, say, the occurrence of infection waves [9, 17] and the effectiveness of containment measures [11].

It should also be emphasized that the `ModInterv` app, by directly implementing mathematical models, provides information about the epidemic dynamics that cannot be obtained neither from a mere visual inspection of the raw empirical curve nor by using only its moving-average smoothed version. Another important aspect of our app is that it makes mathematical analysis of epidemic data freely accessible on the internet to any interested user. Our software was primarily designed to process COVID-19 data, but owing to its generality it can be easily applied to other epidemics, old and new—a possibility that will be explored in the future.

The paper is organized as follows. In Sec. 2 we discuss some general properties of epidemic curves, with emphasis on their different dynamical regimes for epidemics with multiple waves of infections. The mathematical models implemented in the `ModInterv` application to deal with multiwave curves are presented in Sec. 3. A detailed description of the backend structure of the app is presented in Sec. 4, where the several `Python` modules used by the app are discussed. This section also contains a brief description of the LOWESS regression algorithm implemented in the app to identify the number of waves in a given empirical curve. In Sec. 5 we give an overview of the general features of the software and discuss how the user can interact with these features. The user interface is divided into two sections, one for Countries and the other for States and Cities in Brazil and the USA, and the functioning of each section is discussed in detail. Illustrative examples (for Slovakia, Netherlands, and South Africa up to June 2, 2022; and the USA up to July 31, 2021) are given in Sec. 6, where a detailed guide to generate and customize the output graphs is also presented. Finally, the main conclusions of the paper are summarized in Sec. 7.

## 2. Epidemic curves and their dynamical stages

Here we discuss some general properties of typical epidemic curves and their different dynamical growth stages. Our focus will be on epidemic curves with multiple waves. (For a detailed description of single-wave curves, see, e.g., Ref. [24].) Typically, each wave of infection displays three general dynamical regions, namely: i) a period of rapid, accelerated growth at the beginning; ii) an intermediate region where the curve grows approximately linearly in time; and iii) a late growth phase when the curve “bends away” from the linear profile and tends to a saturation plateau. A better, more refined characterization of these three growth phases can be given in terms of acceleration regimes, as described below.

To make the discussion general, let us assume that the full epidemic curve has *N ≥*1 waves. A schematic of a cumulative epidemic curve with two waves of infection (*N* = 2) is shown in Fig. 1a, with the corresponding velocity and acceleration curves shown in Figs. 1b and 1c, respectively. The beginning of each wave (say, the *k*-th one, for *k* = 1, …, *N*) corresponds to a regime of *increasing acceleration*, when the acceleration grows from nearly zero at the end of the previous wave (or the onset of the first wave) and reaches a maximum value at some time, denoted by 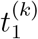 and indicated in Fig. 1 by orange vertical lines. After this time, the epidemic wave enters an intermediate phase characterized by two acceleration regimes: i) first we have a regime of *decreasing acceleration*, where the acceleration decreases from the maximum at 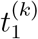 and reaches zero at a time 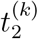, indicated by yellow vertical lines in Fig. 1. The point of zero acceleration (within a given wave) corresponds to the peak of the velocity curve, as indicated by yellow circles in Figs. 1b and 1c. After this point, the acceleration becomes negative and increases in magnitude (see Fig. 1c), thus starting the regime of *increasing deceleration*, which ends at the time, 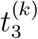, indicated by green vertical lines in Fig. 1, when the deceleration is maximum (the acceleration is minimum). After 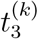, there begins the late growth phase when the deceleration starts to decrease and tends to approach zero towards the end of the wave.

**Figure 1:**
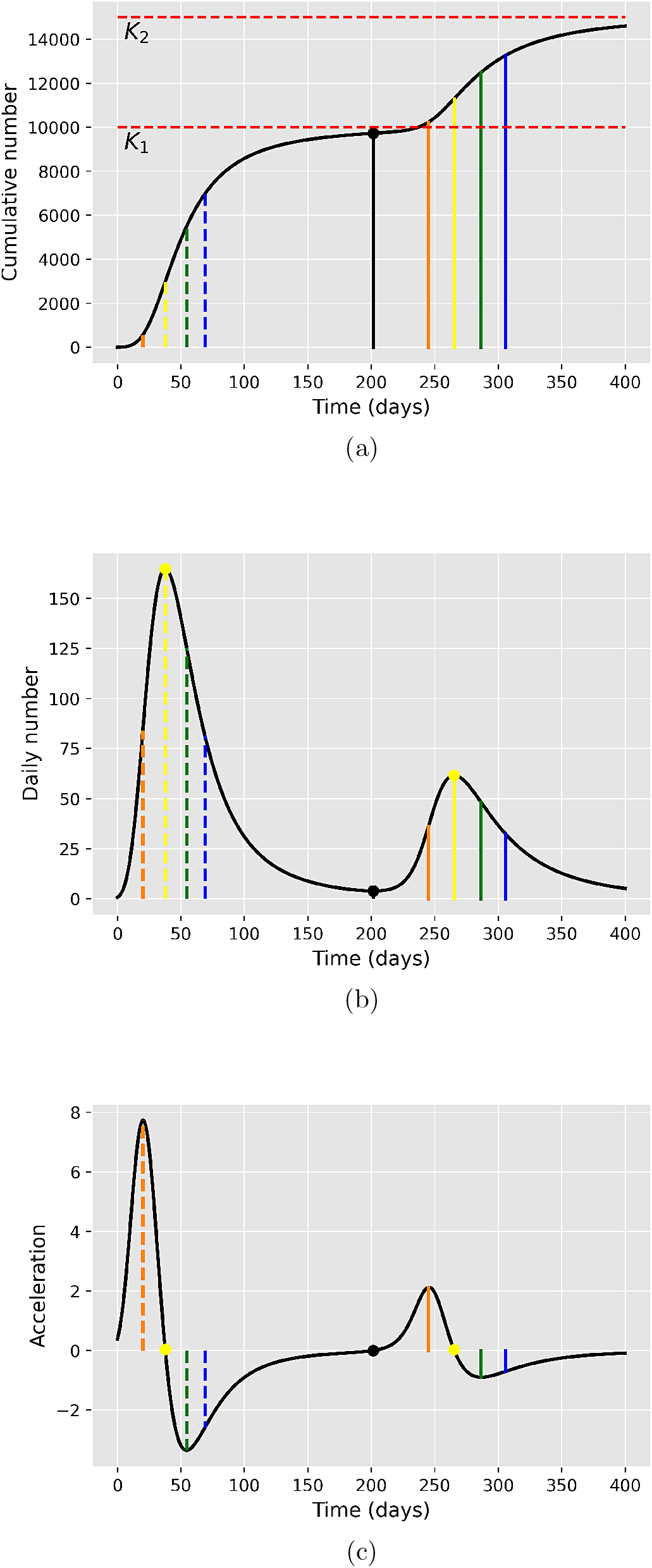
(a) Schematic of a cumulative epidemic curve with two waves of infection, illustrating the five main acceleration regimes in each wave, as separated by the colored dashed (solid) vertical lines for the first (last) wave. The black dot and the solid black line descending from it indicate the beginning of the second wave. (b) Velocity and (c) acceleration curves, corresponding to the first and second derivatives, respectively, of the cumulative curve shown in (a). The yellow circles in (b) indicate the wave peaks.

This late growth phase, with *decreasing deceleration*, is of particular interest because it indicates that the epidemic wave has well passed its “peak” (yellow circles in Fig. 1b) and is now entering its final phase. Because of its relevance for monitoring the possibly approaching end of an epidemic wave, it is convenient to divide this regime of decreasing deceleration into two distinct dynamical stages, according to whether the rate of change of the acceleration, known as jerk, is increasing or decreasing. In the first of such stages, the jerk increases from zero (at time 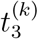) and reaches its maximum at some time which we denote by 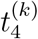 (indicated by blue vertical lines in Fig. 1). As the effect of the positive jerk is to start to bend the curve away from its near-linear profile seen in the intermediate phase [25], we shall refer to this regime of decreasing deceleration and increasing jerk as indicating a *transition to saturation*. The second stage of the late growth phase, which starts at 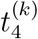, corresponds to a regime of decreasing deceleration and decreasing jerk, which will be referred to as the *saturation* of the epidemic wave. In this final stage the first three derivatives of the growth profile are all decreasing functions of time (in absolute values), indicating that the epidemic curve is approaching a saturation plateau (i.e., the wave’s end).

Summarizing, the characteristic points that we shall use to classify the dynamical stages of an epidemic wave are as follows: i) the point 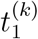 of maximum acceleration; ii) the point 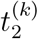 of maximum velocity (zero acceleration); iii) the point 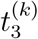 of minimum acceleration (or maximum deceleration); and iv) the point 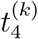 of maximum jerk in the deceleration phase. The end of the *k*-th wave (and beginning of the next if *k < N*) is defined as the point of minimum velocity (zero acceleration) after the wave peak; see black dot in Fig. 1b. The plateau that the cumulative curve would reach at the end of the *k*-th wave (if there were no subsequent wave) is denoted by *K*_*k*_; see Fig. 1a for an example in the case of two waves (*N* = 2), where *K*_1_ and *K*_2_ are indicated by the red dashed horizontal lines.

Knowledge of the points 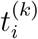 allows us not only to identify the various dynamical regimes of each wave but also to classify the ‘current stage’ of an ongoing epidemic (i.e., up to the final time considered). More specifically, by comparing the position of the last data point of the empirical curve, to be referred as the ‘current time’ *t*_*f*_, with the characteristic points 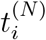 of the *last wave*, the `ModInterv` software (to be discussed later) classifies the current stage of a given epidemic curve according to the following five epidemic stages:

1. Increasing acceleration: 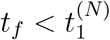.

2. Decreasing acceleration: 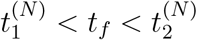.

3. Increasing deceleration: 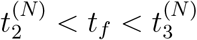.

4. Transition to saturation: 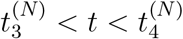.

5. Saturation: 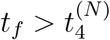.

In order to determine the characteristic points 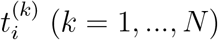, one first needs to fit the empirical data with an appropriate mathematical model. In our software `ModInterv`, the model fits are implemented automatically, i.e., without human assistance, for epidemic curves with an arbitrary number *N* of waves; see Sec. 3. After performing the fit to the chosen data, the software displays in the legend box of the output graph the current stage of the epidemic (among the 5 stages above), together with colored vertical lines indicating the points 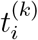 for each wave (according to the color convention above).

As will be discussed below, the generic *N* -wave model used by the app `ModInterv` does not have an exact solution, so that the calculation of the characteristic points 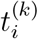 has to be performed numerically. More specifically, after the numerical fit is done, an interpolation of the corresponding theoretical curve generated by the fitted model is made using splines. The location of the maxima and minima of the second and third derivatives of the spline curve are computed, thus determining the position of the four vertical lines for each wave, as illustrated in Fig. 1.

## 3. Mathematical models implemented in the app

As already mentioned, in order to determine the dynamical regimes of a given epidemic curve, it is first necessary to adjust a mathematical model to the empirical data. The `ModInterv` app implements a class of generalized logistic growth models that can be applied to epidemic curves with any number *N* of waves. An automated algorithm — based on the LOWESS regression analysis — is first applied to determine the number *N* of waves present in the data, as will be described in Sec. 4.4. Once *N* is known, the appropriate *N* -wave model is applied. In this section, we give a brief description of the models implemented by `ModInterv`.

The cumulative number *C*(*t*) of cases/deaths in the `ModInterv` is generally described by an extended logistic model, known as the beta logistic model (BLM), defined by the following ordinary differential equation [25, 22]:

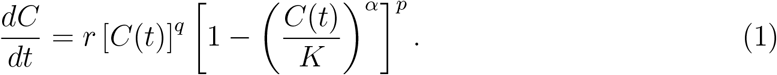

In order to deal with epidemic curves with multiple waves, we assume that all model parameters depend on time, that is, *r* = *r*(*t*), *q* = *q*(*t*), *α* = *α*(*t*), *p* = *p*(*t*), and *K* = *K*(*t*), so as to reflect the changes in the underlying epidemiological parameters during each wave. The specific time-dependency of the model parameters is given by the following multistep logistic-like function [23]:

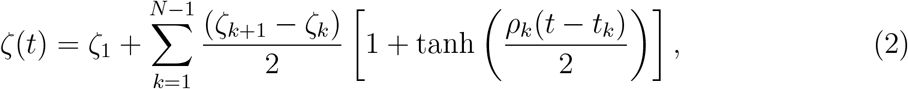

where *ζ*_*k*_ (*k* = 1, …, *N*) represents the corresponding parameter values during the *k*-th wave and *N* is the total number waves. A schematic of the generic parameter *ζ*(*t*), as defined in (2), is shown in Fig. 2 for the case of two waves (*N* = 2). The parameter *t*_*k*_ in (2) determines the transition time between the *k*-th and the (*k* + 1)-th waves; whereas the parameter *ρ*_*k*_ characterises how rapid this transition is, so that the larger the parameter *ρ*_*k*_ the quicker the transition towards the next wave. Note that the characteristic time scales *t*_*k*_ and the corresponding transition rates *ρ*_*k*_ are the same for all parameters. This is justified because an overall change in the epidemic dynamics, brought about, say, by implementation/relaxation of control measures or by a change in the population behavior (or both), is expected to affect simultaneously all epidemiological parameters.

**Figure 2:**
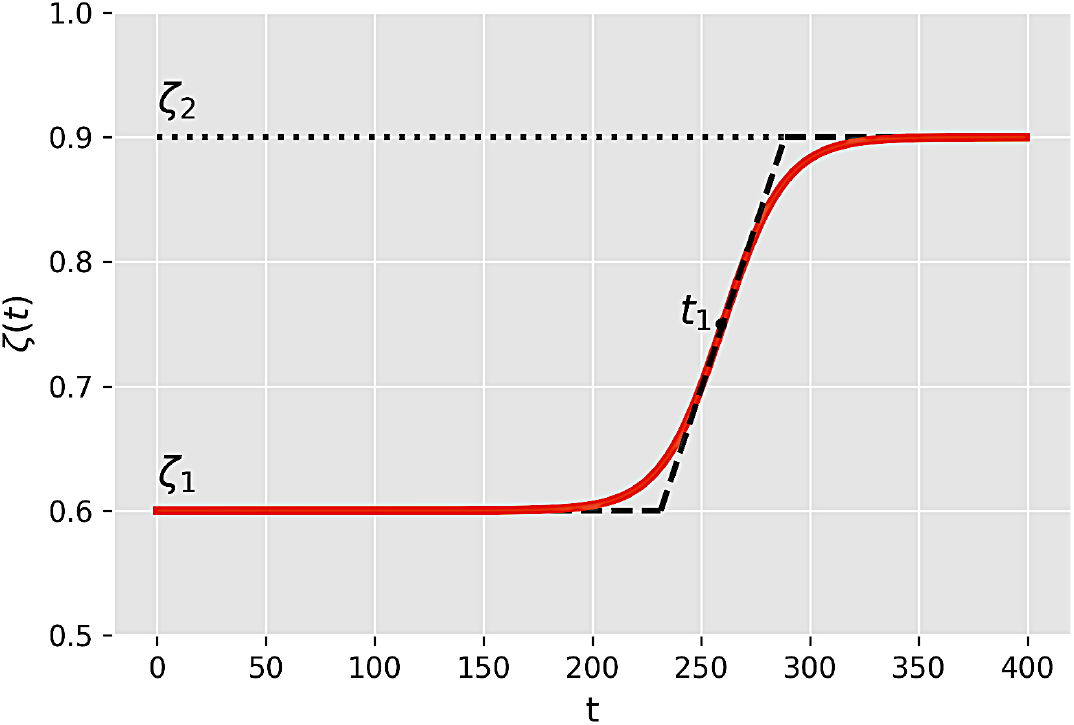
Time dependence of the generic parameter *ζ*(*t*) of the *N* -wave model, as defined by the logistic function given in (2), for the case of *N* = 2 (two waves). The dashed line represents the linear approximation to the logistic function, where the inclined straight line meets the upper and lower horizontal lines at the points *t*_1_ *±* 2*/ρ*_1_, where *t*_1_ and *ρ*_1_ are respectively the transition time and rate between the two plateaus.

For single-wave curves (i.e., *N* = 1), the BLM parameters *{r, q, α, p, K}* are constants in time, see Eq. (2), and can be interpreted as follows [25]: *r* is the growth rate at the early stage; *q* controls the initial growth profile and allows to interpolate from linear growth (*q* = 0) to sub-exponential growth (*q <* 1) to purely exponential growth (*q* = 1); the exponent *p* controls the late-time growth rate, with *p >* 1 implying a polynomial decay of the daily curve after the peak, whereas *p* = 1 yields a fast exponential decay; the exponent *α* controls the degree of asymmetry with respect to the symmetric S-shape of the standard logistic curve (which is recovered for *q* = *p* = *α* = 1); and, finally, *K* is the final size of the epidemic, meaning that *C*(*t*) = *K*, for *t* → ∞. In the case of constant parameters, the BLM admits an analytic solution [25], which is convenient for numerical purposes. The BLM is a versatile model which reproduces other simpler growth models as particular cases [22, 25]. For a detailed discussion of how the `ModInterv` implements the BLM (and some of its particular cases) for one-wave curves, the reader is referred to Ref. [24]. Here our main focus of interest concerns the application of the BLM to multiwave epidemic curves.

The *N* -wave BLM, as given by Eqs. (1) and (2), has a total of 7*N* −2 free parameters— namely, *N* plateaus for each of the five parameter functions (*r*(*t*), *q*(*t*), *α*(*t*), *p*(*t*), and *K*(*t*)) and 2(*N −*1) values for the transition times, *t*_*k*_, and the rates of transition, *ρ*_*k*_, between each successive wave—which need to be fitted from the data. Implementing such a fitting procedure in a way that it operates without human assistance (i.e., for generic input data) poses two main technical challenges. First, one needs a robust algorithm for deciding how many waves there are in a given empirical curve. As discussed in Sec. 4.4, in our app `ModInterv` we have implemented a LOWESS smoothing regression algorithm that allows us to determine the exact number *N* of epidemic waves present in the data. The second technical difficulty is to devise an automated fitting procedure that can handle the large number of free parameters (as *N* increases) without incurring in excessive over-fitting. To minimize the risk of over-fitting we have implemented several control measures in the `ModInterv`, as discussed in Sec. 4.5.

One important point to note here is that once a mathematical model is adjusted to the empirical data for a given location, several important information concerning the epidemic evolution in that location can be extracted from the model, such as the characteristic points 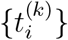 separating the distinct acceleration regimes, which in turn allows the app to determine the current stage of the epidemic curve according to the classification scheme discussed in Sec. 2. The number of waves present in the epidemic curve and its current stage are shown in the legend box of the output plot. Additional relevant information, such as the dates of the peaks of the daily curve as well as the starting dates of each subsequent wave are also given in the output plot, as will be described in Sec. 5. But before going into that, let us present the general backend structure of the `ModInterv` software.

## 4. Software system: backend structure

In this section we will explain in detail the workflow of the software behind the app `ModInterv` and all the tools used in its implementation. A simplified schematics of the functioning of the app is shown in Fig. 3. A detailed description of the different app components is given below.

**Figure 3:**
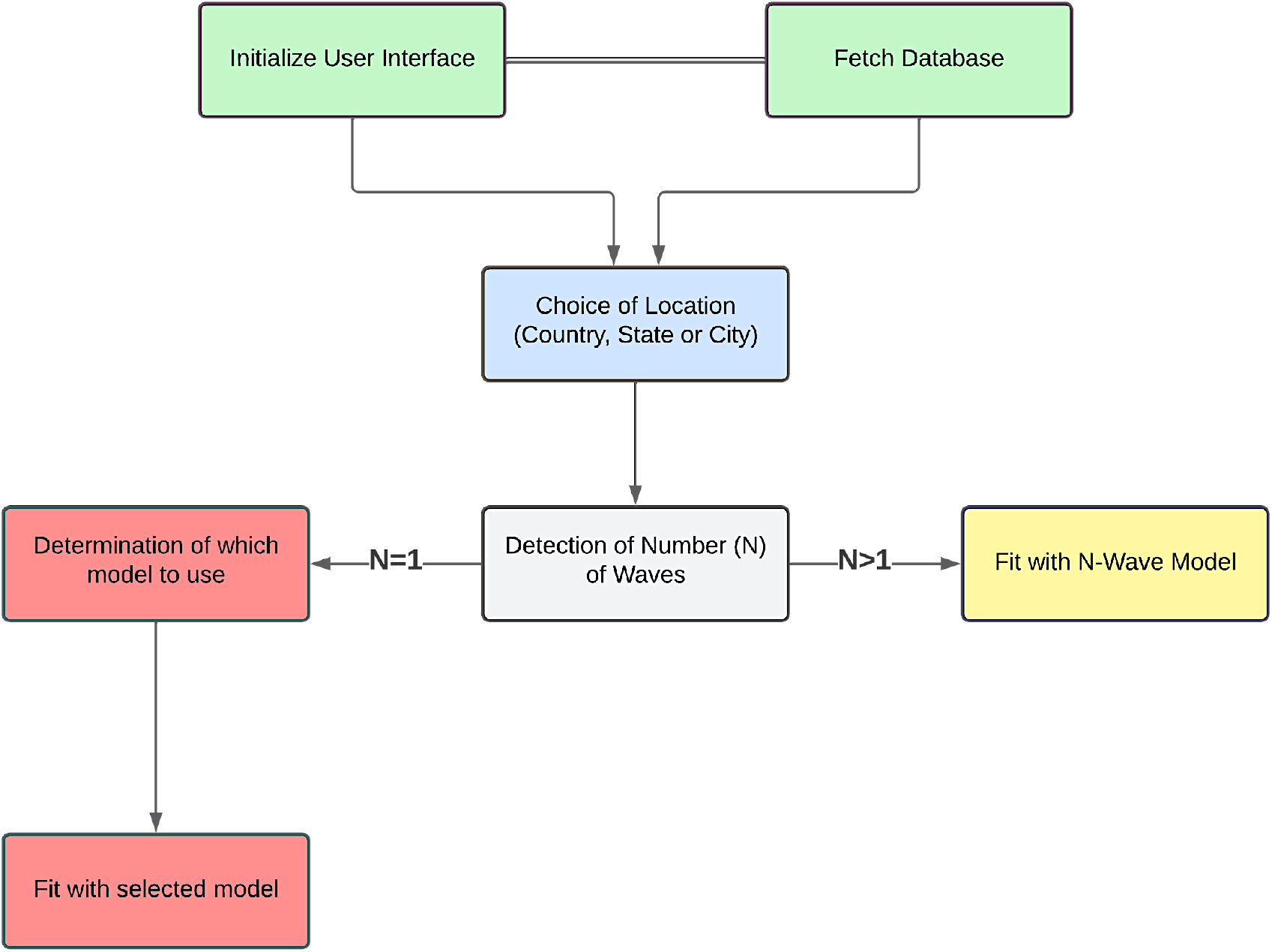
Simplified schematic description of the workflow of the software behind the app `ModInterv`.

### 4.1. Initialization

When the app is accessed (via browser or Android App), the user interface starts to load and the square buttons that appear along the page will read “*Loading widgets*…”; after a few seconds the text in the square buttons will change to “*Show Widgets*”. When the user clicks any of these buttons, the text in all of them changes to “*Initializing widgets*…”; and after a short while a brief introductory text about the app appears. After a few more seconds, the rest of the app will be loaded and ready to be used, as explained below.

### 4.2. User interface

The user interface of the App `ModInterv` was designed using objects called *Widgets*, which are implemented in the `Python` module `ipywidgets`. This module offers a vast miscellaneous of *Widgets* to accomplish the most diverse tasks. In the development of the app `ModInterv` we have used only five types of *Widgets*, namely: HTML, Button, IntSlider, Dropdown and Checkbox.

The HTML *Widgets* is used to display dynamic text throughout the app, avoiding the whole page to be reloaded when the user changes the language (from Portuguese to English or vice-versa). The other four *Widgets* are used to receive inputs from the user, such as the type of the epidemic curve to be fitted (i.e., whether Cases or Deaths), the Country, State or City chosen, the number of days in the epidemic curve to be considered in the fit, among others. Illustrations of each widget are shown in figure 4.

**Figure 4:**
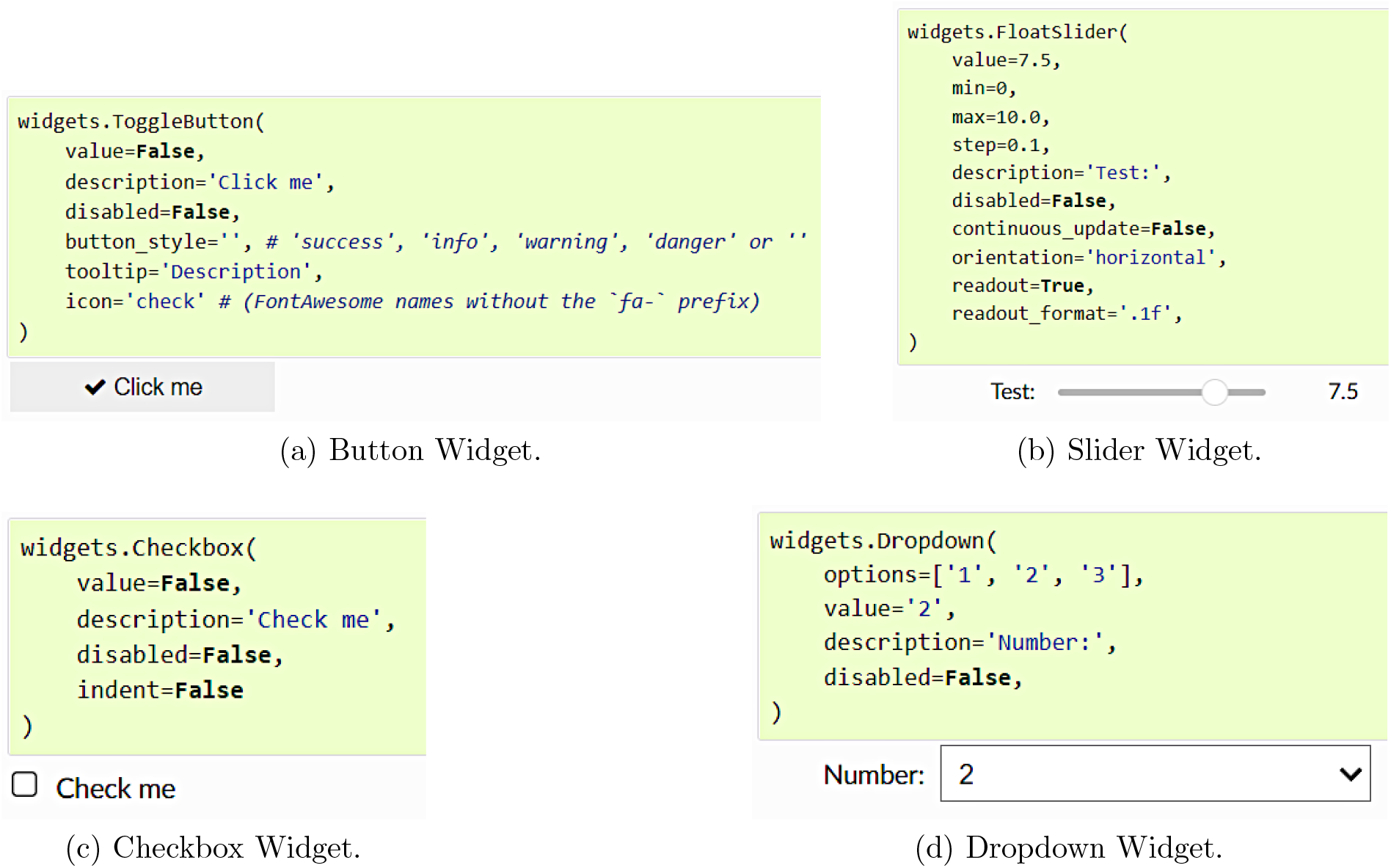
Illustrations of some of the Widgets from the `iPywidgets` module used in the app

### 4.3. Data acquisition

The COVID-19 data for Countries used in the `ModInterv` are obtained from the database made publicly available by the Johns Hopkins University [12], which lists in automated fashion the number of the confirmed cases and deaths for each country in their database. The data used for States and Cities in Brazil were obtained from the GitHub database maintained by Wesley Cota [8] at the Federal University of Viçosa, which is also updated daily. Each time the app is initialized, both databases are accessed and the data for all Countries, States and Cities become available for the user.

In both databases the cumulative counts for cases and deaths attributed to COVID-19 are made available in `.csv` format, which are automatically read by our software once a type of data (Cases/Deaths) and location (Country/State or City in Brazil or US) is chosen. Up-to-date daily cumulative counts for cases and deaths are reported by the two databases above. To obtain daily epidemic curves we subtract consecutive values of the the corresponding cumulative curves. The empirical data are processed by the app as they are stored in the databases and no pre-processing (cleaning) of the data is required. It may happen sometimes that the health authorities of a given country make corrections in their official data, by either discarding deaths wrongly attributed to COVID-19 or by adding a large number of COVID-19 deaths not previously tabulated. In such cases one may see negative daily counts or relatively large upward jumps in the daily curve. In general, however, these extraneous fluctuations have little impact on the overall analysis carried out by the app, as all numerical fits are performed on cumulative curves (for which such corrections are relatively smaller).

### 4.4. Procedure for detecting epidemic waves

The classification of curves that in principle may feature an *a priori* unknown number epidemic waves is quite a challenge to be implemented automatically via an algorithm based solely on mathematical and computational techniques. Although a trained human eye can easily identify when there are multiple waves of infections in a given empirical curve, this is non-trivial task to be performed using purely mathematical quantities, such as derivatives, averages, etc, because the empirical data often present considerable short-scale fluctuations (noise) in addition to longer-scale variations (waves).

In order to determine the number *N* of waves in a given data set, the app `ModInterv` implements an algorithm based on the LOWESS [6] (Locally Weighted Scatterplot Smoothing) smoothing procedure, a method built upon local linear and nonlinear least squares regression. Besides the LOWESS method, we also use a signal analysis `Python` module to detect local maxima and minima, from which the number of epidemic waves present in the data can be estimated. More specifically, the procedure for wave detection works as follows. Once an epidemic data set is selected by the user, by choosing a location and type of data (cases or deaths), the corresponding daily curve is obtained from the successive increments of the chosen cumulative curve. Then, the LOWESS smoothing method is applied to the empirical daily curve, after which a signal analysis `Python` module is used to detect the local maxima and minima of the LOWESS-smoothed curve. Finally, the number *N* of waves in the data is determined as the number of minima plus one.

### 4.5. Numerical fitting

In all numerical fits performed by the `ModInterv`, the app employs the Levenberg-Marquardt (LM) algorithm to solve the non-linear least square optimization problem, as implemented in the `lmfit` module for the `Python` language, which has a built-in routine for estimating the errors of the fitted parameters via the covariance matrix [20].

The numerical fitting of the generic *N* -wave model is quite challenging (especially for large *N*) because each model parameter in the set {*r*(*t*), *q*(*t*), *α*(*t*), *p*(*t*), *K*(*t*)} varies in time according to a logistic-like function with *N* steps (plateaus), one for each wave. In addition to the values of these *N* plateaus, the logistic function contains 2(*N −* 1) additional parameters, namely: the transition times, *t*_*i*_, and the rates of transition, *ρ*_*i*_, between each successive wave; so that in total there are 7*N −*2 free parameters to be fitted from the data. Because of the large number of fitting parameters, special care is required to minimize over-fitting issues. For example, the fit results are sensitive to the initial guesses for the free parameters, which must be carefully chosen so as to yield reliable estimates for the final parameters. With this aim, our fitting procedure for the *N* -wave model (with *N >* 1) is performed in *N* steps, as follows.

In the first step, we give an initial educated guess for the possible location of the first transition time, *t*_1_, between the first and second waves. We then fit the data up to this time with the one-wave model, as given by its analytical solution [25]. The initial guesses for the parameters {*r*_1_, *q*_1_, *α*_1_, *p*_1_, *K*_1_} in this first step are chosen arbitrarily in their ranges of definition; and similar choice is adopted for the initial guesses of the ‘last’ plateau in each step.

Next, we select an initial guess for the second transition time, *t*_2_, between the second and third wave, and fit the data up to this time with a two-wave model, where the parameters found in this first step are used as initial guesses for the first plateau in the two-wave model fit. If the data has only two waves (i.e., *N* = 2) this completes the fit. Otherwise (if *N >* 2), the returned values of the parameters obtained from the two-wave (partial) fit, namely {*r*_*i*_, *q*_*i*_, *α*_*i*_, *p*_*i*_, *K*_*i*_}, for *i* = 1, 2, together with *t*_1_ and *ρ*_1_, are used as initial guesses for the third step, where the data is fitted up to a time *t*_3_ with a three-wave model; and so on until we fit the entire data with the full *N* -wave model. Note that in this way the initial guesses for the set of intermediate plateau parameters {*r*_*i*_, *q*_*i*_, *α*_*i*_, *p*_*i*_, *K*_*i*_}, *i* = 1, …, *N −*1, are estimated in successive partial fits with increasing number of waves. This procedure tends to reduce the sensitivity of the results of the full (final) fit on the parameter initial guesses, thus circumventing excessive over-fitting.

In addition, we have taken extra steps to further minimize over-fitting issues. In Ref. [25] we have found that imposing certain restrictions on the range of the (constant) BLM parameters {*r, q, α, p}* helps to reduce over-fitting when dealing with single-wave curves. Here we adopt the same range restrictions for the plateau parameters *r*_*k*_, *q*_*k*_, *α*_*k*_, and *p*_*k*_, for *k* = 1, …, *N*, as those adopted in the single-wave case, namely: *p*_*k*_ *≥* 1, 0 *< q*_*k*_ *≤* 1, 0 *< α*_*k*_ *≤* 1, and 0 *< r*_*k*_ *<* 1. Furthermore, the parameters *α*_*k*_, for *k* = 2, …, *N* are kept fixed at unity, since we observed that letting them free tends to cause over-fitting. This can be explained by the fact that the parameter *α* is connected with the asymmetry of the daily curve around a peak [25]. But estimating these parameters for the successive waves is less reliable, and so we prefer to set *α*_*k*_ = 1 from the second wave on, so as to reduce over-fitting. Furthermore, the parameter *p*_*N*_ for the last wave is kept fixed at unity. We recall that the parameter *p* controls how fast the daily curve decays after a peak [25], but for an estimate of *p*_*N*_ to be reliable the last wave needs to have a well-formed tail, which is not always the case for a still evolving epidemic. So we decided to set *p*_*N*_ = 1 to avoid possible over-fitting issues with this parameter.

### 4.6. Classification of the epidemic dynamical stage

Once the numerical fit has been performed for a chosen empirical curve and the parameters of the corresponding growth model are determined, the app then computes the points 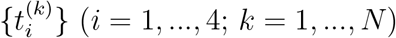 that characterize the distinct acceleration regimes of the curve. These points are computed numerically after interpolating the theoretical curve with a spline and determining the corresponding zeros of the second, third, and fourth derivatives; see Sec. 2. Recall that each set of characteristic points 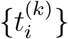 divides the respective *k*-th wave into five acceleration regimes, namely: i) increasing acceleration; ii) decreasing acceleration; iii) increasing deceleration; iv) transition to saturation; and v) saturation. As discussed in Sec. 2, by comparing the final time, *t*_*f*_, of the last point of the empirical data (assumed to be the ‘current time’) with the computed characteristic points 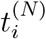 of the last wave, the app then determines the current stage of the epidemic in that location. The corresponding stage for the chosen epidemic curve is then informed by the app in the legend box of the output graph, which shows the empirical data and the fitted curve, together with additional relevant information obtained from the fit, as will be discussed in Sec. 5.

### 4.7. Creating an interactive user interface

The code for the `ModInterv` app was written using `iPython` notebooks, a versatile cloud computing environment that allows for developing codes whithout the need to install all dependencies, as one would usually do when compiling codes directly from one’s own computer. Unfortunately, `iPython` notebooks are not well suited for sharing with public users, because it does not have a user-friendly interface. Furthermore, in order for the users to be able to compile the code and access the app, every one of them would need to have editing permission, which could lead to undesirable results.

Thus, in order to generate an interactive webpage that could be accessed by the users with a customizable, user-friendly interface and with the code running in a backend separate from the interface, we have used `nbinteract`, a `Python` module that creates a static HTML that allows widgets to remains interactive by using `Binder` servers as the computational backend. The HTML file produced then has to be hosted in a server to be publicly accessible on the internet. The HTML page thus produced for the `ModInterv` app can be currently accessed via the address http://fisica.ufpr.br/modinterv. Below we shall explain the contents of the app, as accessed via this homepage. [There is also an Android version of the `ModInterv` available at the Play Store (https://play.google.com/store/apps/details?id=com.tanxe.COVID_19).]

## 5. Software system: output graphs and interaction with user

As already mentioned, the `ModInterv` app allows the user to select the type of epidemic curves (i.e., either Cases or Deaths) to be analyzed as well the location of interest. To help the user choose the location of interest, the app is structured into two main sections: i) Countries and ii) States and Cities in Brazil and the US. The app also allows the user to choose how the output graphs are displayed on screen as well as to generate figures to be downloaded. A brief explanation of the app features is given below.

### 5.1. Countries section

In the first section of the app, the user can analyze the curves of cases and deaths for countries. After the user selects from corresponding dropdown menus the type of `Data` (Deaths or Cases) and the chosen `Country`, a preview with the respective accumulated curve (red circles) is generated for the total number of deaths/cases as a function of time, measured in days from the first death/case.

After clicking the button `Perform fit`, a text box with the message “*Computing*…” will appear, and soon after a new graph with the model fit (black curve) superimposed on the data (red circles) will be displayed below the original preview plot. The name of the selected country is shown in the plot title, together with the date up to which the empirical data was considered, while the model that best fits the data is indicated in the legend box. The plot also shows colored vertical lines corresponding to the characteristic points *t*_*i*_ that define the five acceleration regimes of an epidemic wave, as explained in Sec. 2. Also shown in the fit plot are black dots on the fitted curve, from which it is drawn solid black vertical lines to mark the separation between each successive wave. In the legend box of the plot the app also shows complementary information, such as i) the calendar date of the first case/death; ii) the starting dates of the each wave; and iii) the current dynamical stage of the epidemic, according to the classification scheme presented in Sec. 2.

Besides the information displayed on the plot of the fitted cumulative curve, the user has several additional options to further analyze the results of the fitting process. First, checking the checkbox `Check to display/hide the daily curve` shows the empirical daily curve (red circles) together with the theoretical daily curve (black curve), where the latter corresponds simply to the time derivative of the mathematical fit for the cumulative curve. Second, checking the checkbox `Check to display the parameters of the fit` will produce a table showing the estimated parameters of the best fitted model together with their errors. Third, moving the slider `Time range` gives a short term prediction, ranging from 7 up to 28 days after the last data point, for the number of extra cases/deaths during the selected period ahead. The total cumulative number of cases/deaths at the end of this period is also shown. Fourth, the user has the option to choose the scale on both the x-Axis (horizontal axis) and y-Axis (vertical axis) between the linear and logarithmic scales. Finally, clicking on the buttons Generate figure file will generate links for downloading the figure files for the cumulative and daily curves as well as for the table with the respective fitting parameters, where the formats `png` and `eps` are supported.

To see a new type of data or choose a different country, the user only needs to select the desired options from the corresponding dropdown menus. After a new selection is made, the button `Perform fit` has to be clicked again to produce the fitted plot for the newly chosen empirical data.

### 5.2. Section of States and Cities in Brazil and the US

In the second section of the app, the user can analyze the death and case curves for states and cities in Brazil and states and counties in the United States (US). First, the user must select the desired Country (Brazil or US), after which the Type of data (Deaths or Cases) has to be selected. The user then needs to select the type of Region (States or Cities) and the desired state or, if the option Cities was selected for Region, the desired city (in Brazil) or county (in the US). Again, a preview graph will be generated with the selected data for the chosen location. The rest of the procedure is as explained in the Country section.

## 6. Illustrative examples

In this section we demonstrate the `ModInterv` application for locations with multiple waves of infections and discuss each step of the process. First, we consider cases where the full empirical curve is used in the analysis, i.e., the app fits the chosen curve up to the most recent data available for the selected location. Subsequently, we briefly discuss an extra feature of the `ModInterv` that allows the user to fit the data only up to a selected earlier date.

### 6.1. Full curve fits

Here we show illustrative examples produced by the `ModInterv` as applied to full multiwave epidemic curves. To reproduce plots similar to the examples shown here, the user can go to the first section of the app and select from the `Data` menu the type of epidemic curve to be analyzed (in this case we chose Deaths) and the desired `Country`. First we show the case of Slovakia, which is one of the few countries that has had (so far) only two waves of COVID-19 (most countries have had three or more waves). Once the Country is selected, a preview of the chosen epidemic curve is displayed, as shown in Fig. 5. After that, the user can proceed to click the button `Perform fit` shown at the bottom of the preview plot, after which a small bar written “*Computing*…” will appear and, soon after, the result of the numerical fit will be shown, as illustrated in Fig. 6. (Fits with larger number of waves may take a little longer.)

**Figure 5:**
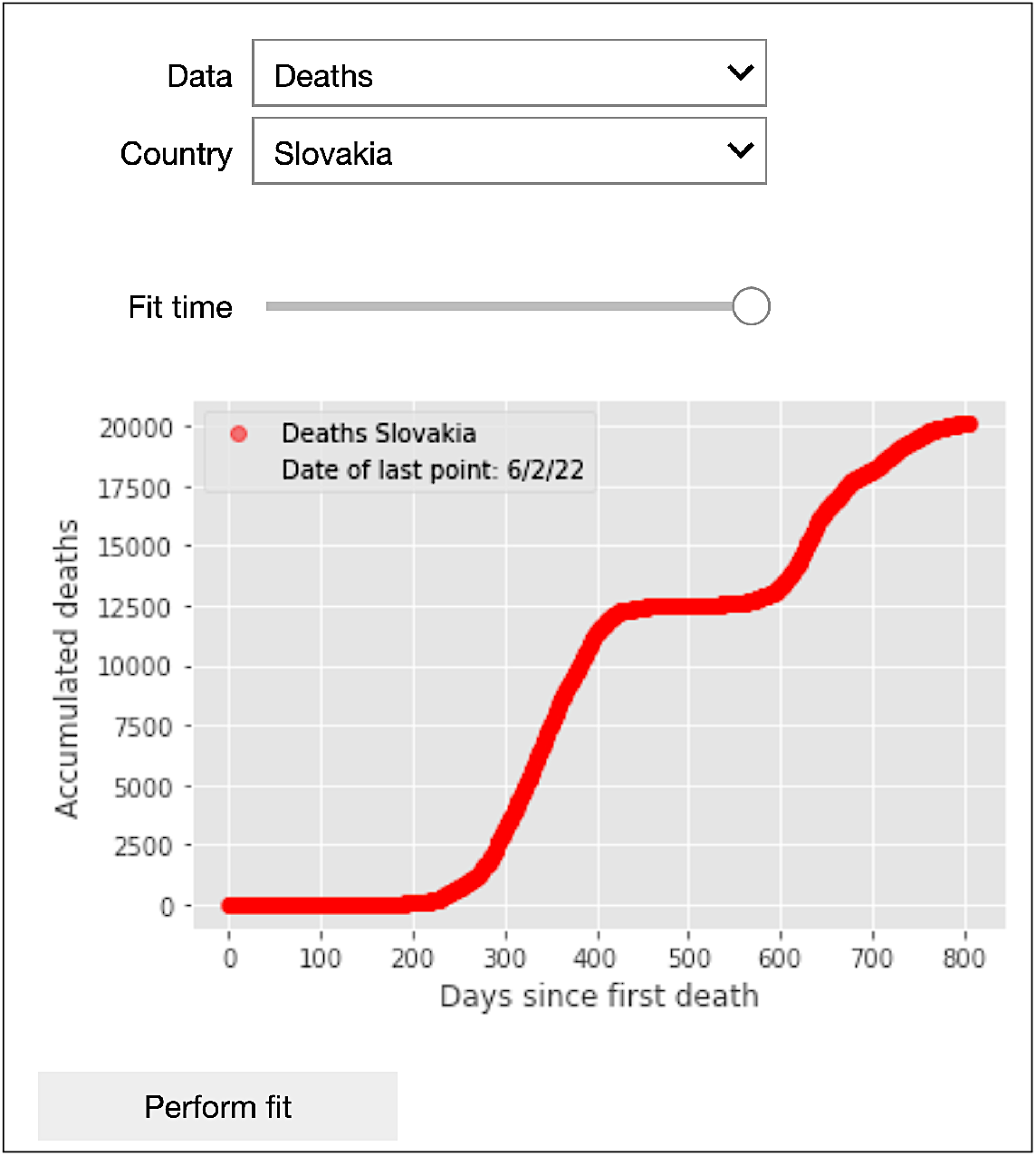
Preview output graph of the app `ModInterv` after selection of the type of Data (Deaths) and Country (Slovakia).

**Figure 6:**
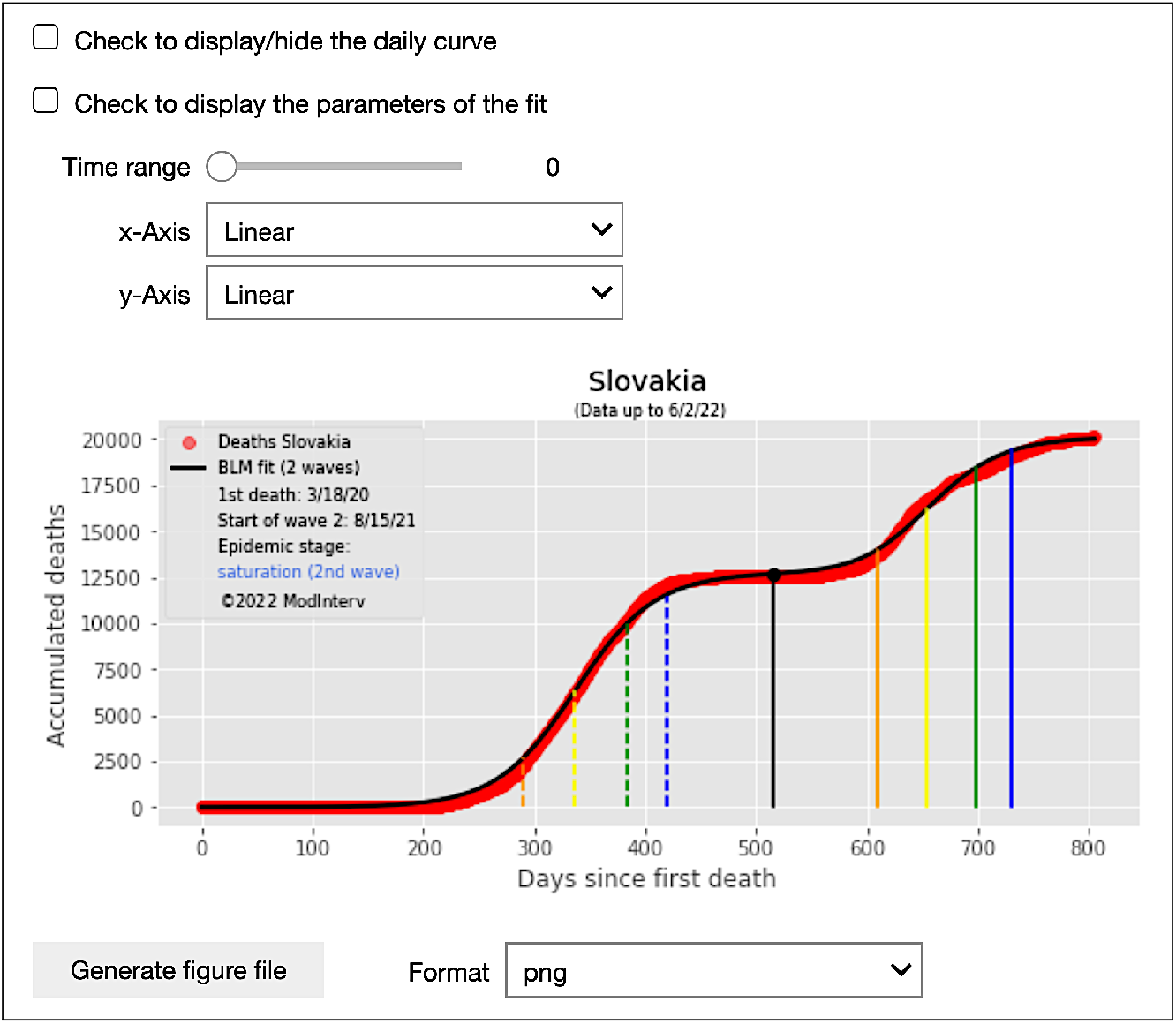
Output of the numerical fit of the epidemic curve (Deaths) for the selected country (Slovakia).

Just above the output plot of the numerical fit, there are two checkboxes written `Check to display/hide the daily curve` and `Check to display the parameters of the fit`, respectively; see Fig. 6. Checking the first checkbox will produce the corresponding daily epidemic curves (both empirical and theoretical), as shown in Fig. 7. In this figure, the red dots correspond to the daily number of deaths and the black line is the time derivative of the theoretical cumulative curve shown in Fig. 6. Also shown in the plot of the daily curves are the locations (yellow dots) of the peaks of each wave, with their respective calendar dates being given in the legend box; see Fig. 7.

**Figure 7:**
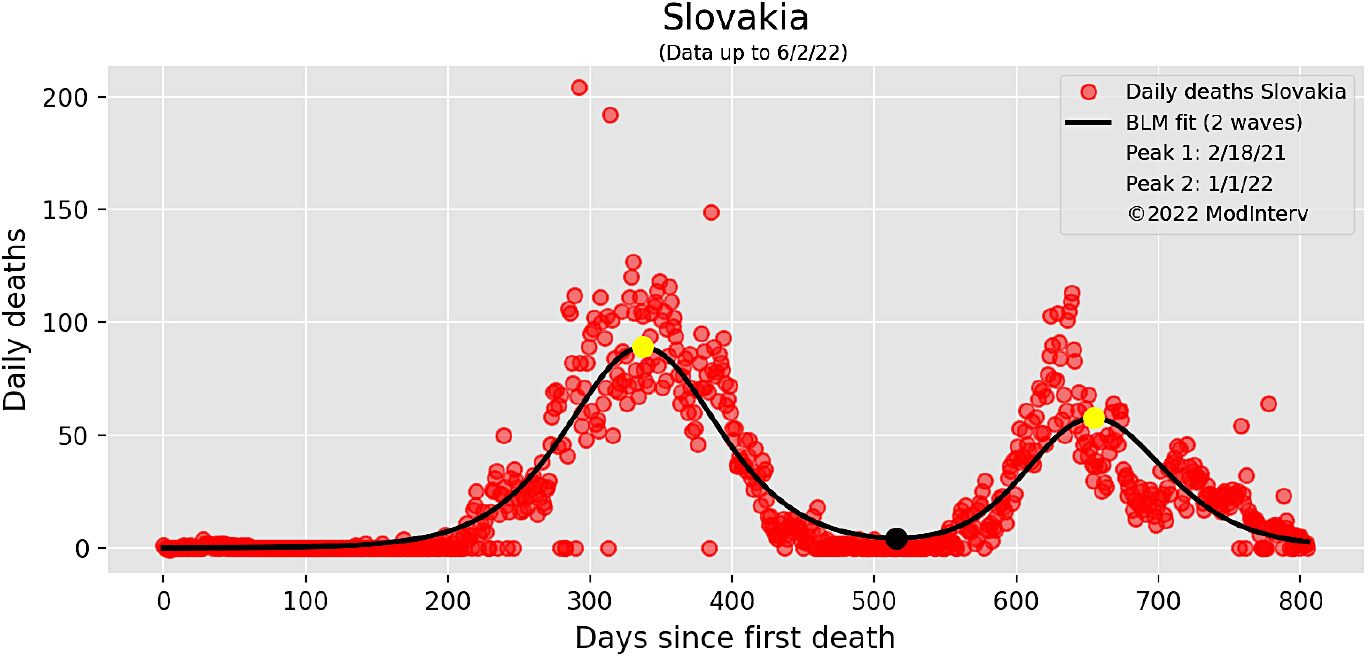
Daily curves (empirical and theoretical) corresponding to the cumulative curves shown in Fig. 6.

Checking the second checkbox renders a table on the screen with the values of the fitted parameters and their respective errors, as illustrated in Fig. 8. Also shown in this table are the values of the characteristic times, *t*_*i*_, separating the different acceleration regimes (*i* = 1, ‥, 4) for each *k*-th wave (*k* = 1, …, *N*); see Sec. 2.

**Figure 8:**
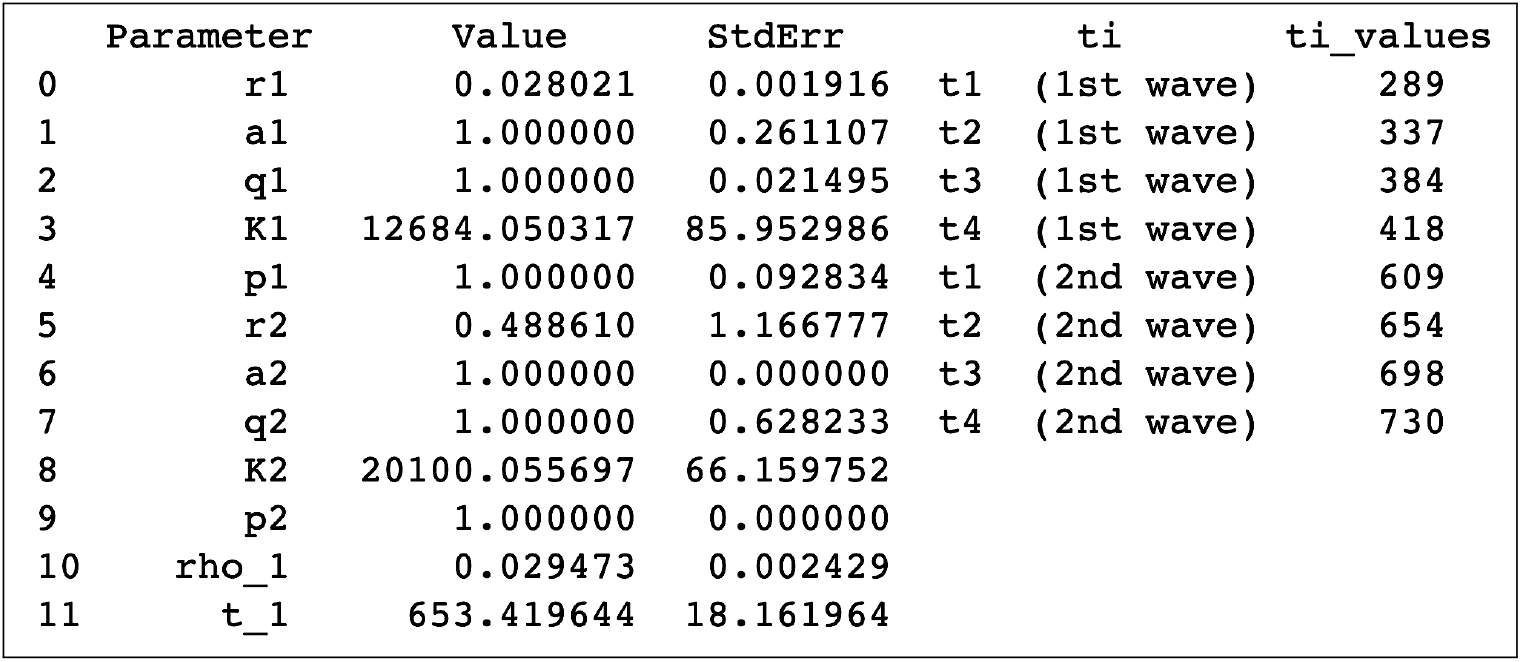
Parameter estimates and respective errors for the numerical fit shown in Fig. 6. Also shown in the table are the value of the characteristic times *t*_*i*_ separating the different acceleration regimes (*i* = 1, …, 4) for each wave.

Note that as seen in Figs. 6 and 7 the theoretical curves are in remarkable agreement with the empirical data, which lends credibility and reliability to the epidemic parameters (such as the beginning, peak, and end of each wave as well as their distinct acceleration regimes) obtained from the mathematical model. We note furthermore that the app allows the user to download higher resolution figures (formats `png` and `eps` are supported) for the cumulative and daily curves as well as for the table with the corresponding fitted parameters. This feature is activated by clicking on the button Generate figure file which appears at the end of each section (Country or States and Cities) of the app; see Fig. 6.

Examples of fits performed by the `ModInterv` for countries with *three* and *four* waves of COVID-19 are shown in Figs. 9 and 10 for the cases of the Netherlands and South Africa, respectively, up to June 2, 2022. Here again one sees that the theoretical fits (which we recall are performed automatedly and without human assistance) are in very good agreement with the empirical data, even for cases of a relatively large number of waves (*N* = 3, 4).

**Figure 9:**
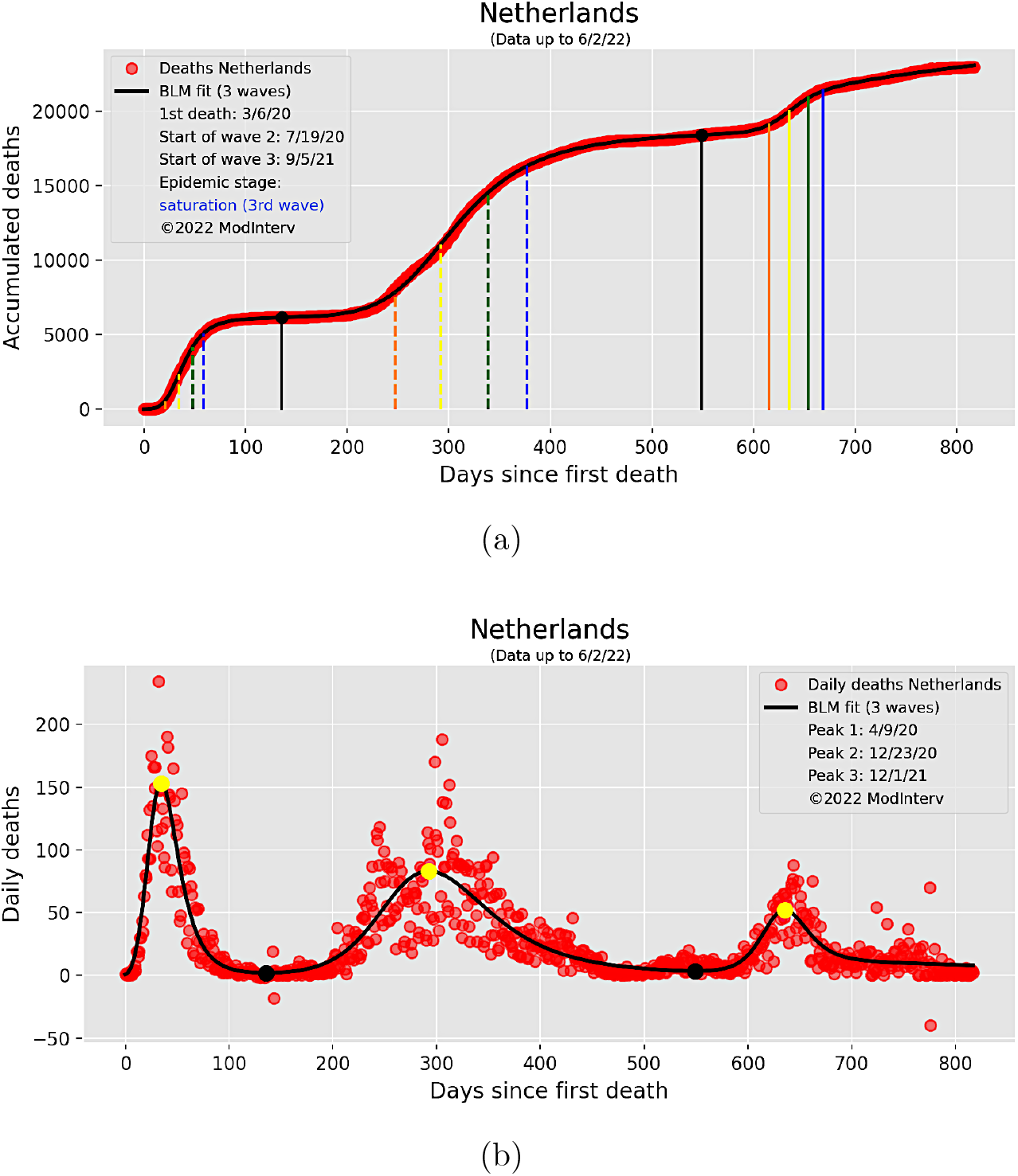
Cumulative (a) and daily (b) curves (empirical data and theoretical fit) for deaths attributed to COVID-19 in the Netherlands up to June 2, 2022.

**Figure 10:**
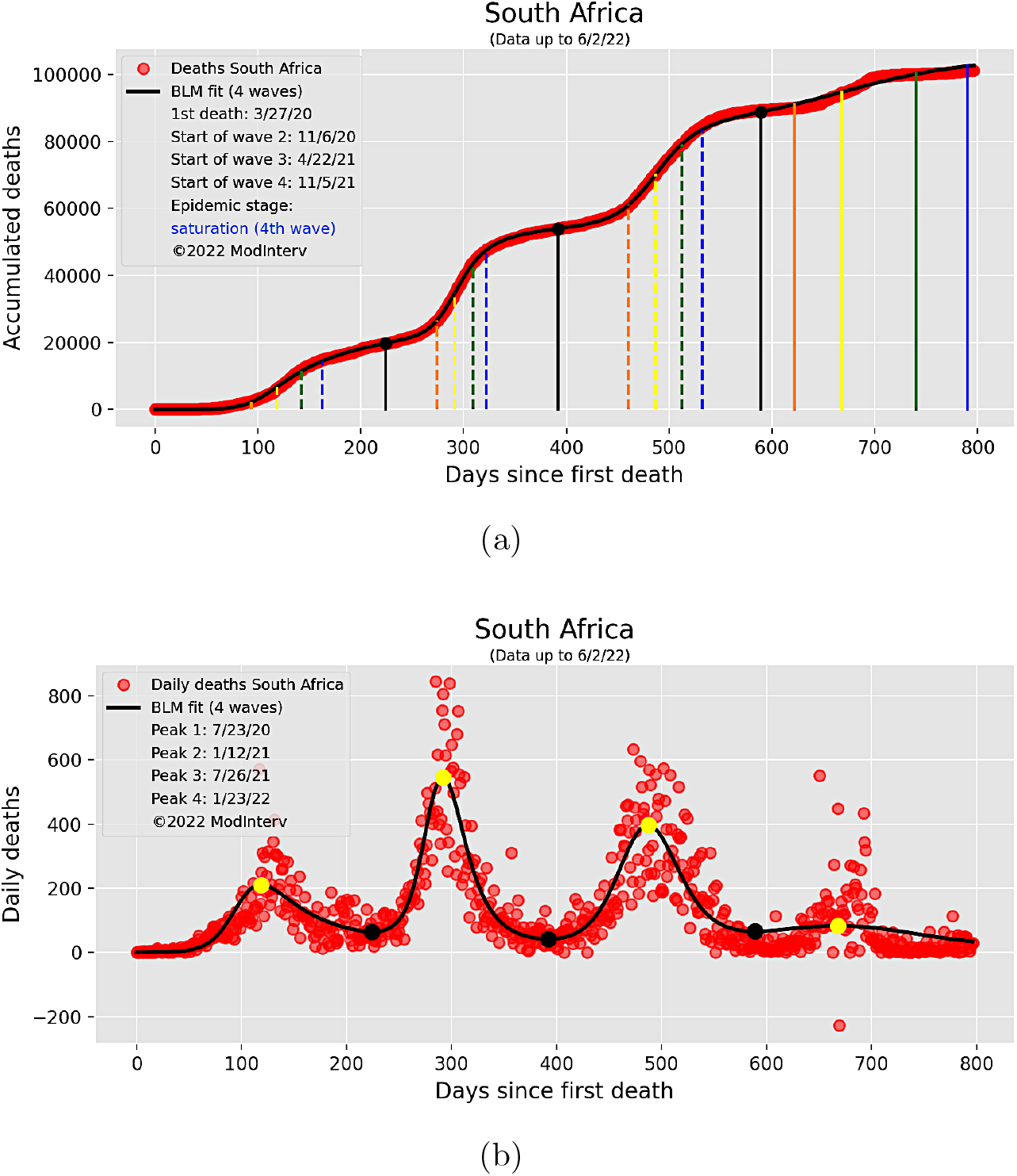
Cumulative (a) and daily (b) curves (empirical data and theoretical fit) for deaths attributed to COVID-19 in South Africa up to June 2, 2022.

### 6.2. Partial fits

The `ModInterv` has an interesting feature that allows the user to truncate the epidemic curve at a past date and perform the numerical fit only up to that date. To exemplify this feature, let us consider the case of the USA up to July 31, 2021. In order to select the desired date, the user only needs to slide backwards the slider written `Fit time`, which appears on top of the preview plot (see Fig. 5 for an example of a preview plot showing the time slider). Once the new (past) date is selected, the preview plot is automatically updated showing the empirical data only up to the selected date, which is shown in the corresponding legend box of the plot as `Date of last point`. Then clicking on the `Perform fit` button will instruct the app to perform the fit only up to that last point, resulting in the plots shown in Fig. 11 for the case of the USA up to July 31, 2021.

**Figure 11:**
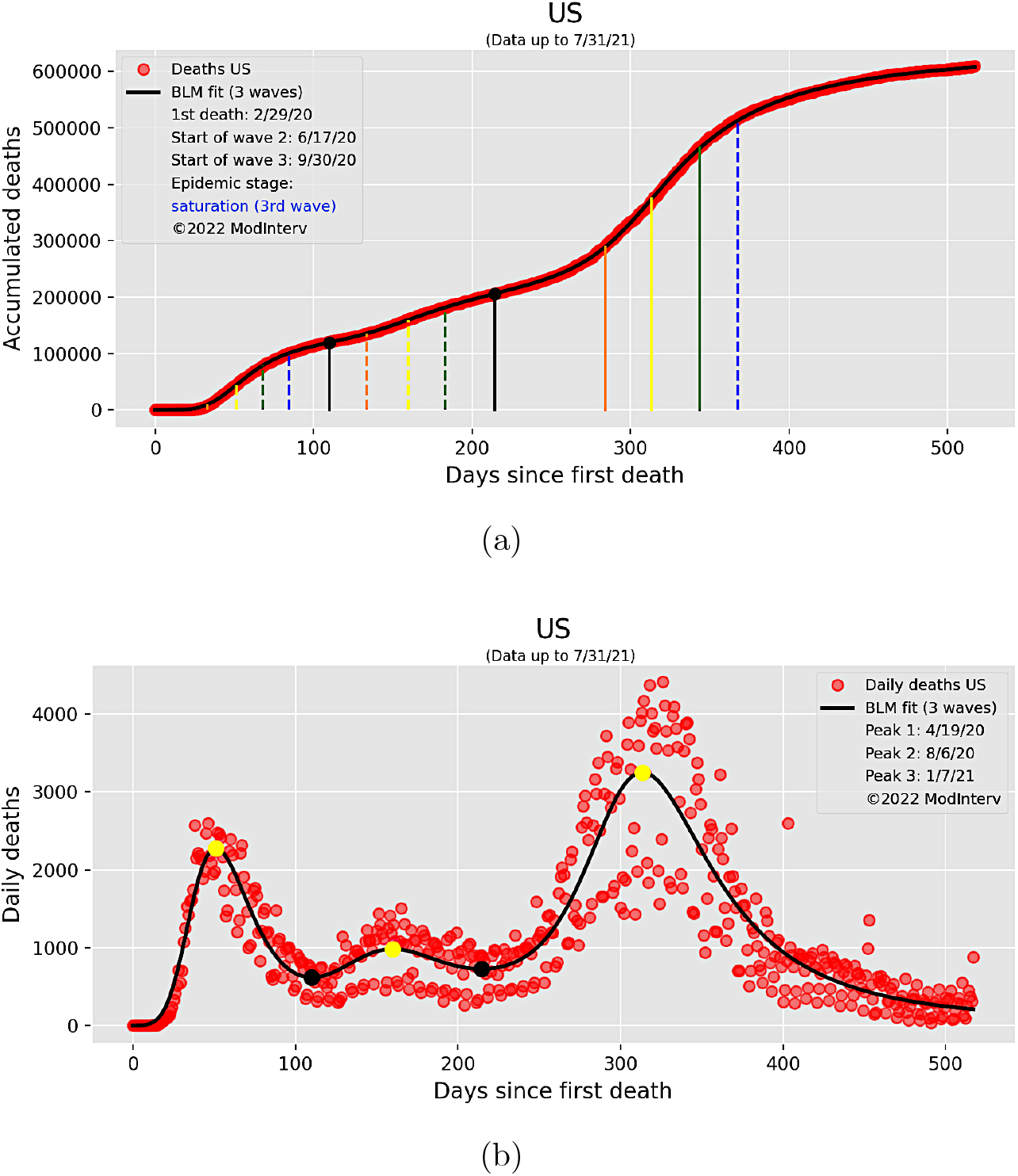
Cumulative (a) and daily (b) curves (empirical data and theoretical fit) for deaths attributed to COVID-19 in the USA up to a time (July 31, 2021) when there were three waves of infection.

## 7. Conclusion

In this paper we have described an automated software application, called `ModInterv`, that enables the user to analyze COVID-19 epidemic curves of cases and deaths for different countries around the world as well as for states and cities in Brazil and the USA. The application uses epidemic data available in public databases. Once a location and type of data (i.e., either cases or deaths) are selected, the app automatically fits the empirical data with a general class of mathematical growth models suitable for epidemic curves with multiple waves. From the best fitted mathematical model, relevant information about the progress of the COVID-19 epidemic in that location can be inferred, such as the dynamical stage of the epidemic in the chosen location.

Our methodology allows for a finer classification scheme of the different growth regimes of an epidemic curve by considering five main acceleration regimes, as explained in Sec. 2. This should be compared, in contrast, with the common way to track epidemics evolution in terms of the effective reproduction number *R*_*t*_, where we recall that *R*_*t*_ *>* 1 (*R*_*t*_ *<* 1) implies that the epidemic is accelerating (decelerating). Although *R*_*t*_ is widely used by epidemiologists and public health authorities, this quantity has nonetheless some drawbacks [2, 1, 24]. For example, as *R*_*t*_ essentially represents a measure of the epidemic acceleration, it cannot by itself distinguish between regimes of increasing or decreasing acceleration (for the same value of the acceleration). In this context, it is therefore useful to have additional tools to obtain a fuller description of the epidemic evolution, say by analyzing also its different acceleration regimes, as implemented in the `ModInterv`.

Furthermore, the classification scheme implemented in our application can provide relevant information to health authorities not only in regard to the current stage of the epidemic in the place of interest but also about its likely evolution in the near future. Such information can help the authorities in their decision making process regarding, say, the implementation or relaxation of non-pharmacological interventions [24]. Moreover, by relying on a general class of flexible growth models, from which the points separating the different acceleration regimes can be easily computed, the classification of a given epidemic curve can be performed automatically by the software (i.e., without human assistance), thus making the method easily accessible to any interested person, without requiring specific mathematical knowledge.

The app also provides additional relevant information about the course of the epidemics which cannot be easily obtained from a visual inspection of neither the raw empirical data nor its moving-average smoothed version. For example, from the fitted mathematical model for a given epidemic curve the app computes more precise estimates not only for the starting and peak dates (points of zero acceleration) for each wave but also the points of maximum acceleration and maximum deceleration. This wealth information, in turn, can be useful for government and health authorities as well as for other researchers, as its allows for a more reliable comparison of the different dynamical stages of the epidemic with the containment measures in place at those particular times. We believe that this type of analysis can help one to understand the underlying reasons for the pattern changes displayed by the epidemic curve.

Our software was primarily designed to process COVID-19 data, but owing to its general structure it could be readily applied to data from other infectious diseases. In this regard, and as an interesting research perspective, one can thus envisage a general purpose platform to analyze data for different diseases. Technically, this would be rather simple to implement. The main requirement is that the relevant empirical data be available in public repositories. Once such repositories are setup, it is only a matter of changing the URL address where to locate the data and the `ModInterv` can run on the new database. This makes our system a relevant informatics tool in the study and control of old and new infectious diseases.

## Data Availability

Johns Hopkins University https://coronavirus.jhu.edu/map.html and GitHub database maintained by Wesley Cota https://github.com/wcota/covid19br

## Acknowledgments

This work was supported in part by the following Brazilian agencies: Conselho Nacional de Desenvolvimento Científico e Tecnológico (CNPq), under Grants No. 312985/2020-7 (GLV), No. 312612/2019-2 (AMSM), No. 305305/2019-0 (RO) and No. 167348/2018-3 (AAB), and Coordenação de Aperfeiçoamento de Pessoal de Nível Superior (CAPES). GLV also acknowledges partial funding from UFPR through the COVID-19/PROIND-2020 Research Program.

